# Spatiotemporal Dynamics of West Nile Virus and Eastern Equine Encephalitis Virus in Georgia Highlight Divergent Ecological Niches for Persistent Transmission Over Two Decades (2001-2025)

**DOI:** 10.64898/2026.07.29.26359272

**Authors:** Thuy Vi Thi Nguyen, Bridget O. Aito-bobadoye, Adem Filali, Elmer W. Gray, Skyler M. Kerr, Rosmarie Kelly

## Abstract

West Nile virus (WNV; Flaviviridae) and Eastern Equine Encephalitis virus (EEEV; Togaviridae) represent the two most significant mosquito-borne zoonoses in the southeastern United States. While both viruses utilize avian amplifying hosts and mosquito vectors, it is unclear if they occur within distinct ecological niches. This study assessed the cumulative effects of both landscape composition and weather variables on the spatial-temporal distribution of WNV and EEEV outbreaks in Georgia over a twenty-four-year period (2001-2025). We used a modeling framework that directly accounts for both spatial and temporal effects but additionally integrates key EO variables (Earth observation). These environmental effects were investigated using Bernoulli generalized linear mixed-effects models (GLMMs) and executed with an integrated nested Laplace approximation (INLA). Our findings revealed disproportional distribution ranges of *Culex quinquefasciatus, Aedes albopictus and Culiseta melanura* in highly urbanized Georgian counties such as Chatham, Dekalb, and Fulton. Results showed that WNV transmission was heavily influenced by urban land cover (Posterior Mean: +0.78, 95% CrI: [0.61,0.95]) but negatively associated with lagged precipitation (Posterior Mean: −0.18, 95% CrI: [−0.29, −0.07]), confirming drought-driven amplification. Conversely, EEEV transmission was strongly influenced by wetland cover (Posterior Mean: +1.12, 95% CrI: [0.94,1.30]) and precipitation (Posterior Mean: +0.51, 95% CrI: [0.39,0.63]). These results underscore the need for improved mosquito surveillance across all Georgian counties in the face of growing vector-borne disease risks.

## Introduction

Mosquito-borne diseases are a significant global health threat, causing about one million deaths annually (LaBeaud et al 2011,Curren et al 2017) . The risk posed by arboviruses and the diseases they cause is escalating as climate change, urbanization, global travel, and trade accelerate shifts in mosquito ecology (CDC 2020, Rosenberg et al 2018, LaDeau 2007, Likos 2016, Ryan 2019). These changes support the range expansion of mosquito species into new geographic regions, resulting in increased disease burden and outbreaks, which are often driven by viral mutations that enhance pathogen transmissibility (Turell 2005, CDC 2018, Barnett 2010). Considering this increasing threat, expanding mosquito surveillance is crucial to support the development of effective prevention or control strategies, which is now an urgent public health priority in the United States.

Several large metropolitan states across the United States are now observing the re-emergence of these mosquito-borne pathogens (Roche 2013). These states face disproportionate arboviral risk due to unique geographic and environmental conditions. Georgia is one such state, being the eighth most populous state in the United States and home to more than 11.3 million residents. Its capital, Atlanta, is a metropolitan area that contains more than half of the state’s population. Georgia’s natural world is equally rich. The Okefenokee swamp is one of North America’s largest black-water wetlands teeming with wildlife and ancient cypress stands. The Chattahoochee and Savannah Rivers carve boundaries and nourish ecosystems across the state that support mosquito breeding. In the north, the Appalachian foothills offer suitable forest ecosystems with more soil types than any other state in the US. All these habitats have created a conducive environment for the introduction of invasive mosquito vectors and sustained transmission of their arboviruses (Althouse et al 2012, Park et al 2016, Lord and Day 2001, Glass 2005, Garrett-Jones 1964, Smith and McKenzie 2004). These arboviruses have notably impacted public health in the state of Georgia, with human cases of WNV reported to be on the rise annually since its introduction in 2001, along with historical outbreaks of EEEV and Californian sero-group viruses (CSG) (Lord and Day 2001, Chaves et al 2011, McMillan et al 2019, Andreadis 2008). Additionally, travel-associated cases of chikungunya, dengue, and malaria have raised concerns in recent times about the potential establishment of emerging or re-emerging arboviruses across the state ( Smith and McKenzie 2004). The year-round presence of key mosquito species vectoring these pathogens could further amplify such threat of pathogen re-emergence and arboviral disease outbreaks.

West Nile virus (WNV) is classified as a neurotropic, mosquito-borne virus belonging to the flavivirus genus (Ryan et al 2019, Turell et al 2005). A mosquito–bird–mosquito transmission cycle is important for its sustenance in nature with *Culex* species broadly recognized as the primary vector and *Aedes albopictus* as a competent bridge vector (Turell et al 2005, CDC 2018) West Nile Virus (WNV) was first reported in the United States in 1999 and the state of Georgia in 2001 respectively, where it has since emerged as the primary cause of arboviral neurological disease (Ryan et al 2019,Turell et al 2005, CDC 2018, Barnett 1964).

Another important arboviral disease is Eastern equine encephalitis (EEE), commonly known as sleeping sickness, which causes inflammation of the brain and spinal cord in equines and humans (Barnett 1964, Roche et al 2013). The primary enzootic mosquito vector of EEEV is *Culiseta melanura*, which develops in and around freshwater hardwood swamp environments. The bridge vectors for EEEV are *Aedes, Coquillettidia,* and *Culex* species for transmission to humans or to horses, which act as incidental dead-end hosts (Turell et al 2005, CDC 2018, Barnett 1964, Roche et al 2013). The disease has a high mortality rate in horses and humans, so is considered one of the most serious mosquito-borne diseases in the United States with cases commonly seen in the southeastern United States (Roche et al 2013).

Although symptoms in humans are relatively rare, the implications can be severe. Approximately 1 in 150 people infected with WNV develops neuroinvasive disease with a mortality rate of approximately 10% (CDC 2018). An average of 8–12 human EEEV cases occur per year in the United States, with an estimated 30% mortality and high rates of permanent neurological damage among survivors (Roche et al 2013). The impacts on un-vaccinated equines from EEEV and WNV are also severe in animals that develop neuroinvasive disease, with fatality rates of 50–70% and 30–40% (CDC 2018), respectively.

Unpredictable shifts in temperature, precipitation, and the frequency of these extreme weather events are altering the ecological balance of many regions in the United States (CDC 2018, Park et al 2016). These changes are creating conditions for more conducive mosquito proliferation and arboviral amplification (Althouse et al, 2012, Park et al 2016). As a result, the interplay between changing climate conditions and the geographic expansion of WNV and EEEV as well as their vectors has become a growing concern with public health implications (Lord and Day 2001, Glass 2005), especially across the United States.

Georgia, located in the southeastern US, represents a critical zone for both WNV and EEEV transmission. This has been attributed to its varied ecological landscapes, supportive climatic conditions, and abundant *Culex, Aedes* and *Culiseta* mosquito populations.

Numerous studies across the United States have identified a range of environmental factors associated with the prevalence of WNV-EEEV infected mosquitoes and associated human cases (LaDeau et al 2007, Likos et al 2016). Temperature and precipitation have consistently emerged as key drivers, with higher temperatures linked to earlier onset and prolonged mosquito seasons, increased infection rates and, potentially, reduced adult mosquito survival during peak summer periods (Garrett-Jones 1964, Smith and McKenzie 2004) Precipitation has shown a more complex relationship with outbreak prevalence. Reduced rainfall, and especially drought conditions, are associated with higher mosquito infection rates and increased WNV and EEEV circulations in some areas of the United States (Roche et al 2013, Park et al 2013). Early-season precipitation has also been linked to delayed mosquito emergence but greater overall abundance later in the season, especially with *Culex* and *Aedes* species (Roche et al 2013, Chaves et al 2011). Ecological and landscape variables, such as proximity to larval habitats, elevation, vegetation indices, land use, and habitat fragmentation, have also been found to influence mosquito abundance and arboviral transmission dynamics (Glass 2005). Additional climatic variables, including wind speed and overwintering conditions, have been shown to affect mosquito survival and resultant WNV-EEEV infection rates (Garrett-Jones 1964). These previous studies reflect the multi-factorial nature of WNV-EEEV transmission dynamics and highlight the increasing influence of climate change on transmission of arboviruses and seasonal dynamics of mosquito vectors.

Due to the lack of effective vaccines for mosquito-borne diseases, mosquito surveillance and control remain the primary approach for arboviral disease prevention. Effective mosquito control depends on robust surveillance programs to continuously provide essential data on mosquito populations, pathogen activity, and environmental factors influencing mosquito dynamics and transmission risk (McMillan et al 2019, Andreadis et al 2014). However, the ecological and spatial complexities in spillover rates of zoonotic arboviruses from mosquito vectors to both humans and livestock pose a huge challenge to developing effective prevention and control strategies. By connecting dense vector monitoring data with widespread availability of multi-scale Earth observation (EO) environmental data (Andreadis et al 2014, Brady et al 2015), arboviral spillovers and potential outbreaks could be more accurately predicted.

Though Georgia has decades of surveillance data, the true geographic distribution of arboviral risk remains insufficiently characterized across all 159 counties. This has been largely due to systematic surveillance bias towards potentially high-risk metropolitan counties due to limited resources and funding cuts. Our surveillance data reflects only the subset of mosquito populations that are successfully collected and submitted for laboratory testing, and therefore do not represent a complete accounting of arboviral activity. Absence of detections were not interpreted as absence of transmission, but rather as a limitation inherent to passive mosquito surveillance across all 159 counties in Georgia. Knowledge gaps of arboviral transmission hot-spots in both urban and peri-urban areas, as well as seasonal dynamics of mosquito vectors and intermittent outbreaks of WNV and EEEV in multiple counties of Georgia, have demonstrated the need to use tools that can leverage dynamic Earth Observation (EO) data.

This study provides a comprehensive spatio-temporal analysis of WNV and EEEV surveillance datasets from 2001 to 2025. Mosquito, avian, and equine data sources were integrated to identify main ecological niches where primary or competent mosquito vectors and arboviral outbreaks predominantly circulate across Georgia’s 159 counties. The spatiotemporal correlations of two zoonotic arboviruses of major human and veterinary health importance in Georgia (WNV EEEV) were investigated utilizing the robust monitoring data set from the Georgia Department of Public (GDPH). By leveraging extensive surveillance data over two decades (2001-2025) from Georgia’s mosquito surveillance programs, we provide useful insights into mosquito population dynamics and emerging transmission hot-spots. We can use this information to develop geographically tailored vector control interventions and pave the way for improved preparedness against potential outbreaks across all Georgia counties.

## Materials and Methods

### Study area

Georgia is in the south-eastern region of the United States, with Atlanta serving as the state capitol. As the sixth most populous U.S. state, Georgia is home to 11.6 million residents within 1706.96 square miles (4428.78 km^2^), averaging 2749 people/mi² (1061 people/km^2^). Georgia encompasses 159 counties that are grouped into six distinct ecological regions: Southeastern plains (n= 64 counties), Piedmont (n=43), Southern coastal Plain (n=31 counties), Southwestern Appalachian (n=8), Mountain (n=15) Blue Ridge (n=1), and Ridge and Valley (n= 8 counties) (**Figure 1**). The average monthly temperatures in Georgia range from 60.7°F (15.9°C) in January to 80.7°F (27.1 °C), and annual rainfall averages about 52 inches (132 cm), peaking from May through October 23. Georgia’s climatic regime, defined by hot summers, mild winters, and extensive wetland systems, all create environmental conditions that not only support high mosquito abundance but also enhance the vector competence of *Culex*, *Culiseta*, and *Aedes* species for arboviral transmission.

**Figure 1:**
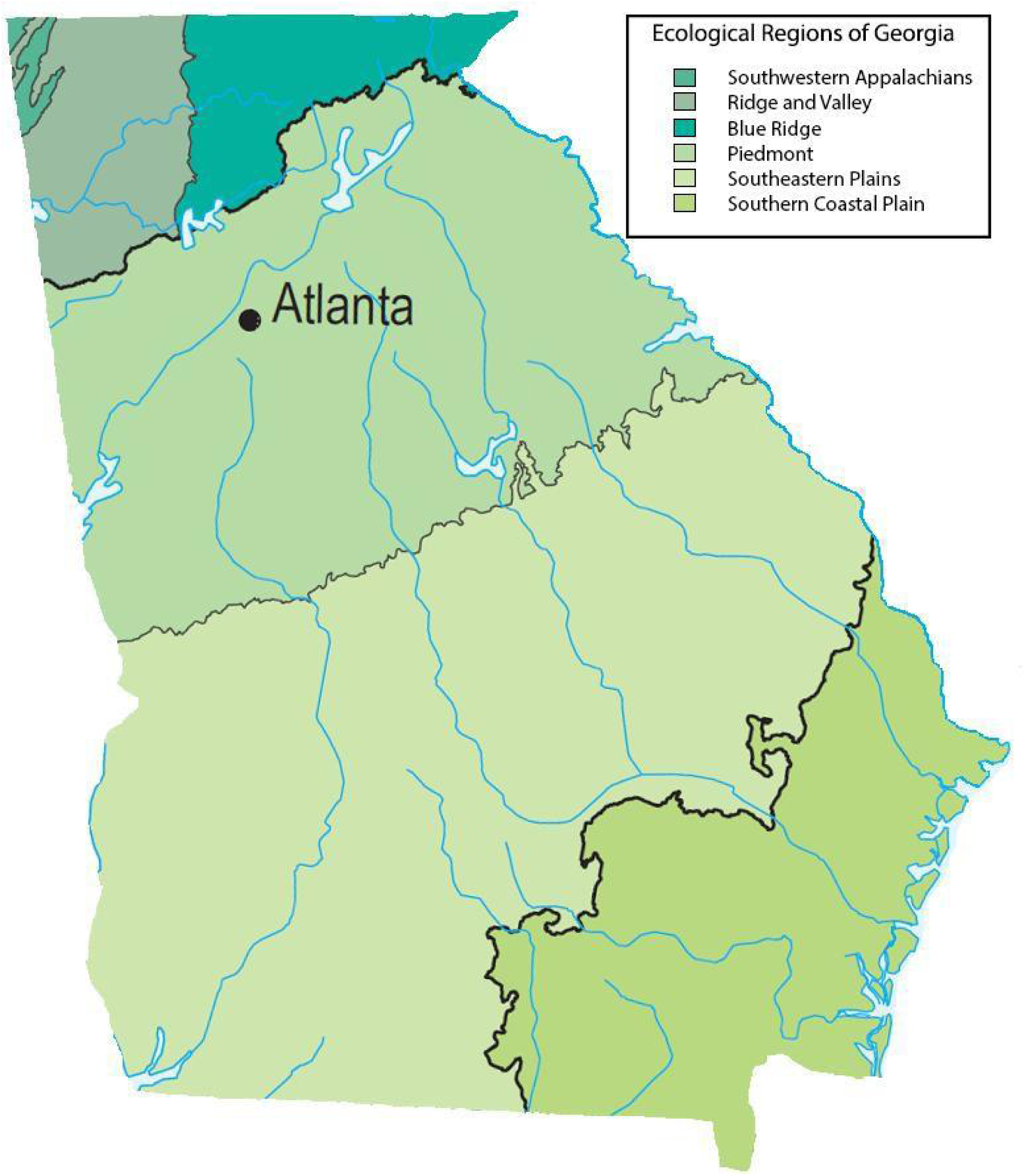
Ecological regions of the state of Georgia.

### Data sources

We obtained adult mosquito surveillance data (2001-2025) from the Georgia Department of Public Health (GDPH). GDPH conducts routine mosquito trapping across the state as part of its vector surveillance program. A preponderance of the sampling was conducted to monitor West Nile virus and eastern equine encephalitis virus circulation across the state. Datasets were checked for completeness, including gaps in surveillance types (dead bird, mosquito and equine surveillance) across all counties from 2001-2025.

Four surveillance datasets were integrated for this analysis. Data included the MIR datasets for all counties comprising of WNV positive pool records; EEEV and WNV case data were confirmed from both equine and dead bird surveillance **(Figure 2)**. Mosquito surveillance data included mosquito species, trap nights, and collection dates. Temperature, precipitation and land cover data were obtained from the National Oceanic and Atmospheric Administration website (https://earthexplorer.usgs.gov).

**Figure 2:**
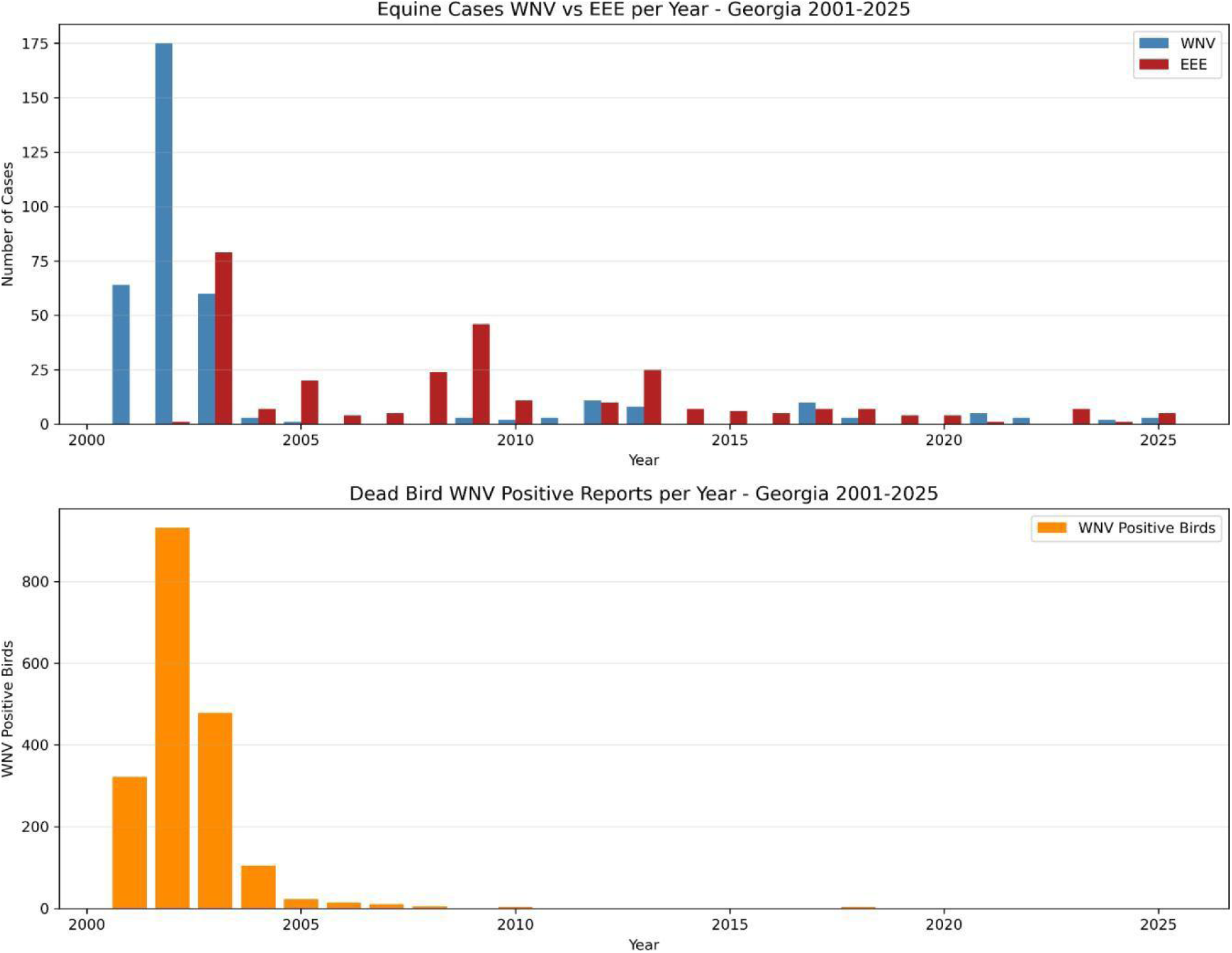
Annual temporal trends in WNV and EEE surveillance across two biological hosts in Georgia (2001–2025), showing equine cases and avian mortality.

### Statistical Analysis

To model the binary presence/absence of both arboviruses across sampled locations, we implemented a Bayesian spatiotemporal Bernoulli Generalized Linear Mixed Model (GLMM) because this framework is well-suited for surveillance datasets where outcomes are discrete and detection’s are sparse. The Bernoulli GLMM accommodates the probabilistic nature of pathogen detection and allows fixed effects (environmental or ecological covariates) to be estimated while accounting for sampling heterogeneity. Spatial variation was modeled using a Matérn Gaussian Random Field through the Stochastic Partial Differential Equation (SPDE) approach, which provides a computationally efficient method for representing spatial auto correlation across irregularly spaced surveillance sites. This is particularly important for mosquito-based surveillance, where sampling locations are unevenly distributed and spatial dependence must be captured.

Temporal auto correlation was incorporated using a first-order auto regressive process, reflecting the biological reality that arboviral detections in one season are influenced by conditions in preceding seasons (e.g., overwintering dynamics, vector population carryover, or persistent enzootic transmission). This structure allows the model to account for serial dependence in annual surveillance data. Models were executed using Integrated Nested Laplace Approximation (INLA) in R-INLA because INLA provides a fast, accurate Bayesian inference for latent Gaussian models and is widely used for spatiotemporal disease surveillance. To ensure robustness of fixed-effect estimates, a logistic regression equivalent was implemented in Python (Version 3.10), following the statistical validation approach described by Seabold and Perktold (2010).

Surveillance effort was measured as the total number of mosquito pools tested per county over the study period (2001-2025). Counties were classified as high-surveilled (>10,000 pools), moderately surveilled (1,000-10,000 pools), and under surveilled (<1,000 pools). This helped to identify counties where low testing volume may mask true arboviral transmission risks.The surveillance categories were defined using thresholds that reflect meaningful differences in sampling intensity and the expected reliability of arboviral detection under varying pool-testing volumes. High-surveilled threshold (>10,000 pools) will identify counties with sustained, high-intensity mosquito testing over multiple years. At this level of sampling, the probability of detecting circulating arboviruses if present is high, and surveillance data are considered robust enough to support fine-scale spatial inference. These counties typically have mature mosquito-control programs, consistent trap deployment, and year-round laboratory capacity. Moderately surveilled (1,000–10,000 pools) threshold will capture counties with regular but not exhaustive surveillance. Testing volumes will therefore be sufficient to detect moderate to high transmission activity. This category reflects the middle tier of operational capacity, where surveillance is informative but still vulnerable to sampling gaps. Nevertheless, under-surveilled counties (<1,000 pools) will be counties below this threshold that have limited mosquito sampling, often due to resource constraints, sparse trapping networks, or intermittent participation in surveillance. For counties at this level, non-detection is more likely to reflect insufficient sampling rather than true absence of arboviral activity, making these counties susceptible to underestimation of transmission risk.

Mosquito abundance was standardized as the number of female mosquitoes of each species collected at a unique trap location in one night (trap-night), averaged over weeks to adjust for variations in collection frequency at each trap location. Non-parametric statistical methods were used to analyze data on mosquito abundance due to non-normal distribution. Two-tailed Kruskal–Wallis tests were conducted to assess differences in mosquito abundance during confirmed peak seasons (May–October) across the study years. Subsequent pairwise comparisons using Dunn’s test with False Discovery Rate (FDR) correction were conducted to determine specific species differences in mosquito population abundance. Statistical significance was set at α= 0.05. These analyses were performed using R (version 4.3.1.27) (R Core Team. 2023).

Species richness, measured as relative abundance, helped to categorize mosquito species according to Heydemann’s classification, from all sampled locations where surveillance was carried out .Species comprising more than 30% of the sample were classified as eudominant. Those species that made up 10 to 30 % of the sample were classified as dominant. Those comprising 5 to10% were classified as subdominant. Those comprising >1 to 5% were classified as rare. Those comprising less than or equal to ≤ 1% were classified as sub-rare.

## Results

### Mosquito Species Composition

A total of 58 mosquito taxa were identified across all entomological surveillance collections dated from 2001-2025 (**Figure 4**). *Culex quinquefasciatus,* commonly known as the southern house mosquito was the eudominant and most abundant mosquito species, comprising 49% of all collected specimens, which is consistent with its well-documented role as the primary WNV vector in southeastern United States. *Culex spp*. (unidentified) represented 13% of collections, *Aedes albopictus,* known as the Asian tiger mosquito, accounted for 8% of all caught specimens. *Aedes albopictus* is a species of growing public health concern given its demonstrated vector competence for multiple arboviruses and its ongoing geographic expansion under projected climate warming scenarios. *Culiseta melanura,* known as the dark tailed mosquito, is confirmed to be the primary vector of EEEV between bird hosts. This species was identified in 2.4% of all trap collections collected predominantly from locations found within the Coastal Plains of Georgia. Despite its low relative abundance on a statewide scale, *Culiseta melanura* is known to breed primarily in acidic, water-filled cavities (crypts) of freshwater hardwood swamps. It has been found to be highly localized to South Georgia, and its low statewide counts further reflects its strict geographic constraint, yet its high vector efficiency within those swamp boundaries explains why EEEV remains highly endemic and persistent in South Georgian counties. Additionally, approximately 7000 *Coquillettidia perturbans* specimens were collected. Due to its similar aggressive feeding patterns on both birds and large mammals (such as horses and humans), this species commonly known as the cattail mosquito is a bridge vector for EEEV and could be responsible for carrying EEEV out of deep swamps and transmitting it to dead-end equine and human hosts (McMillan et al 2019,Turell et al 2005 ).

### Temporal Trends in WNV Transmission

Analysis of 160,294 mosquito pool records across 25 years revealed four distinct WNV transmission peaks (**Figure 3**). The highest recorded MIR was detected in 2001 and 2011 (MIR= 5.2%, 31 positive pools out of 597 tested and 5.2%, 397 positive pools out of 7,622 tested respectively), followed by a sustained period of elevated transmission intensity between 2017 to 2019 (MIR ranging from 4.39% to 4.70%, representing 276, 310, and 243 positive pools respectively). A fourth peak was observed in 2024 (MIR=2.73%, 256 positive pools). These four transmission peaks exhibit an approximate seven-year cyclical pattern, with inter-peak periods characterized by substantially lower MIR values (range 0.42–2.07%). Surveillance (trapping efforts) were shown to be highly localized geographically with Chatham County accounting for 72,582 pools tested, representing the single largest county-level surveillance effort within the data-set (**Figure 4**). In contrast, 57 of 159 Georgia counties (35.8%) recorded fewer than 100 pools tested over the entire 25-year study period, indicating a systematic under-representation of a substantial portion of Georgia counties in surveillance efforts.

**Figure 3:**
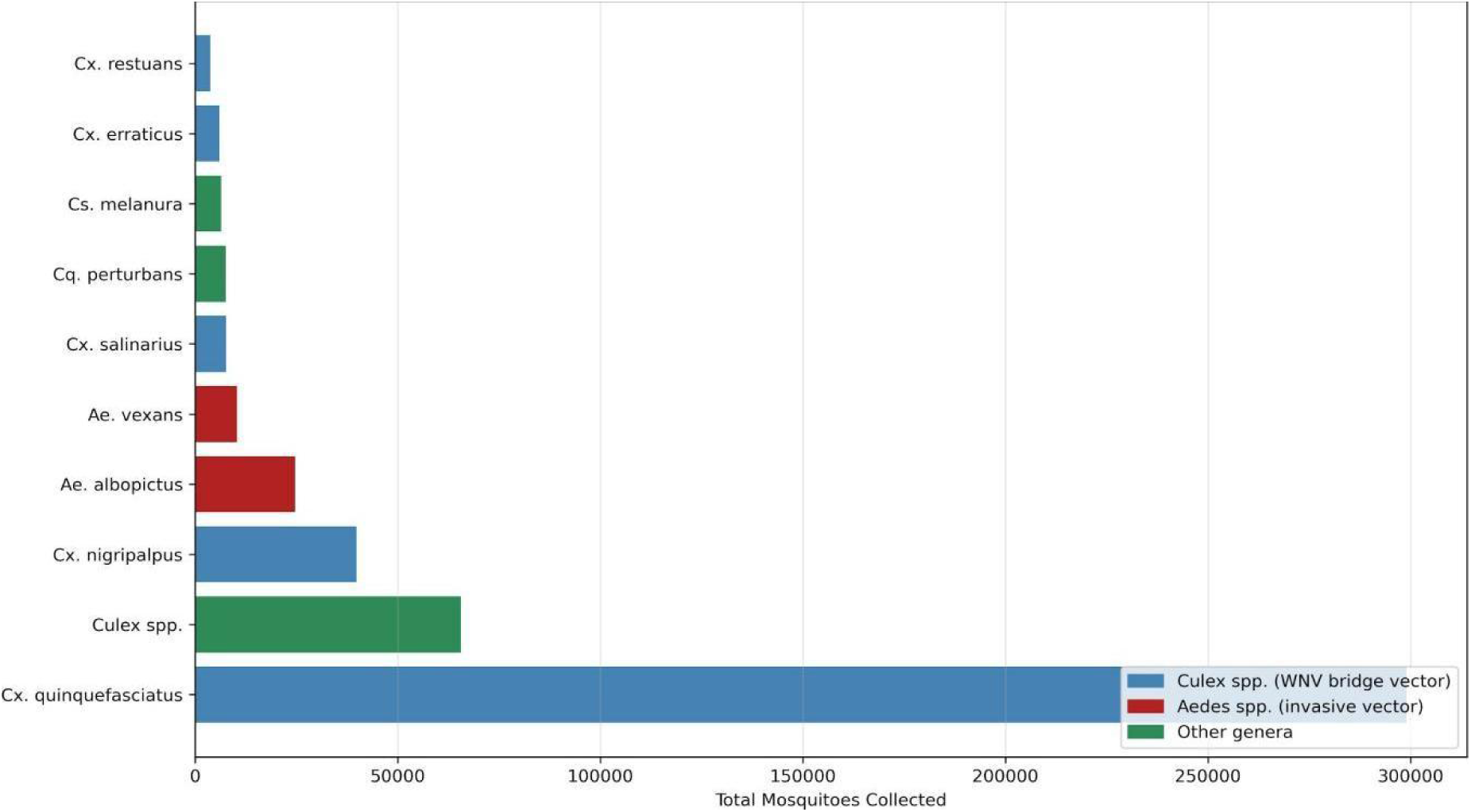
Species composition of mosquito collections in Georgia (2001–2025), showing the top 10 species by total abundance. Blue = *Culex spp*. (WNV vectors), Red = *Aedes spp* (invasive, competent WNV vectors); Green= (Primary EEEV vectors).

**Figure 4:**
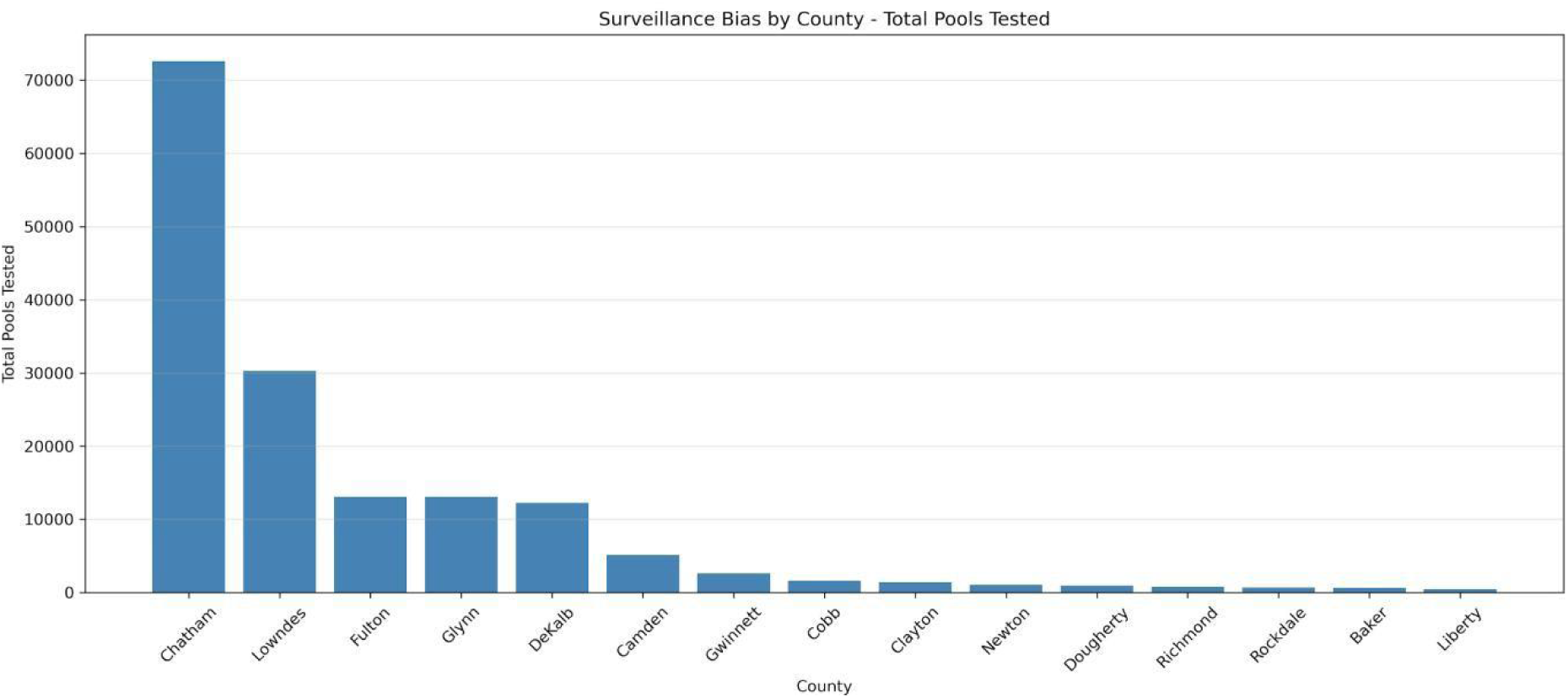
Complete temporal trend of WNV-positive mosquito pools in Georgia revealing trapping effort across all counties (2001–2025).

The fixed-effect posterior estimates from the WNV model revealed this pathogen deeply integrated into human-dominated landscapes and was highly sensitive to acute meteorological anomalies. Urban land cover emerged as the strongest predictor of WNV presence in Georgia (Posterior Mean: +0.78, 95% CrI: [0.61,0.95]) and this strong positive association reflects the ecological preferences of the primary WNV vector, *Culex quinquefasciatus*. In Georgia’s metropolitan centers, aging gray infrastructure, combined sewer overflows (CSOs), and residential artificial containers could create highly eutrophic standing water as breeding sites for this mosquito species. These organically polluted aquatic habitats are highly preferred by *Cx. quinquefasciatus* for oviposition and larval development (Rueda et al 1990).

The model also confirmed a significant negative association between WNV presence and lagged precipitation (prec_lag1) (Posterior Mean: −0.18, 95% CrI: [−0.29,−0.07]). This negative coefficient provides strong empirical support for a drought-driven WNV amplification hypothesis in the southeastern United States. During dry spells, storm-water catch basins and urban streams stagnate, accumulating high concentrations of organic matter that accelerate *Culex* larval growth. Simultaneously, scarce water resources force avian amplifying hosts (passerine birds such as blue jays*, Cyanocitta cristata)* and female *Culex* mosquitoes to congregate by localized stagnant aquatic pools, dramatically accelerating the viral transmission loop while blood feeding.

Lagged temperature (temp_lag1) exhibited a positive effect on WNV presence (Posterior Mean: +0.45, 95% CrI: [0.32,0.58]). Warmer temperatures directly shortens the extrinsic incubation period (EIP) of the virus within the mosquito, allowing vectors to become infectious much faster after an initial blood meal. Additionally, elevated temperatures accelerate larval development rates and increase the biting frequency of adult female mosquitoes (Rosenberg et al 2018, Ryan et al 2019 ).

In stark contrast to WNV, the EEEV model delineated a pathogen strictly constrained by natural, undisturbed wetland or swamp ecosystems and highly dependent on wet hydrological regimes. Wetland cover was the single most powerful predictor of EEEV presence (Posterior Mean: +1.12, 95% CrI: [0.94,1.30]). EEEV is ecologically tied to freshwater hardwood swamps, tupelo-cypress wetlands, and organic-rich soils (Turell et al 2005). These specialized habitats support the development of the primary enzootic vector, *Culiseta melanura,* which breeds in the water-filled cavities (crypts) beneath swamp trees. The negative coefficient for urban land cover in the EEEV model (Posterior Mean: −0.84, 95% CrI: [−1.05,−0.63]) further underscores that EEEV is a rural, forest-interior wetlands pathogen that is highly sensitive to habitat composition and urbanization.

Unlike WNV, EEEV presence was strongly and positively associated with lagged precipitation (prec_lag1) (Posterior Mean: +0.51, 95% CrI: [0.39,0.63]). This variable (lagged precipitation) captures delayed ecological effects over a time period on mosquito populations and arboviral transmission. High water tables and heavy winter/spring rainfall are critical to keeping hardwood swamps flooded, expanding the available larval habitat for *Cs. melanura*. Conversely, dry years will often lead to a rapid decrease of EEEV transmission, as these specialized swamp micro-habitats are prone to drying out completely during drought years. Lagged temperature (temp_lag1) also positively supported EEEV presence (Posterior Mean: +0.62, 95% CrI: [0.48,0.76]). Our results indicated that warmer temperatures could also expand the seasonal transmission window in South eastern Georgia, allowing for earlier spring amplification and extending transmission late into the autumn months.

### Transmission Hot-spot Analysis and Surveillance Bias

County-level MIR analysis revealed a striking inverse relationship between surveillance effort and true transmission risk. DeKalb county emerged as the primary WNV hot-spot with a Minimum Infection Rate (MIR) of 6.67% (816 positive pools out of 12,226 tested), followed by Fulton County (MIR=4.13%, 541 positive pools out of 13,088 tested). Both counties are located within the Atlanta metropolitan Piedmont corridor and are defined by elevated urban density, high impervious surface coverage, and abundant artificial container habitats that support *Culex quinquefasciatus* breeding (**Figure 5**). The Piedmont ecological region exhibited the highest and most persistent WNV transmission risk, with MIR values reaching 22.4% in 2011 and exceeding 10% in multiple years. The Coastal geo-region showed the second highest positivity rates despite constant adulticiding, while the mountain region recorded very minimal WNV activity, a pattern consistent with both limited *Culex* breeding habitat at higher elevations and reduced surveillance intensity in these counties (**Figure 6**).

**Figure 5:**
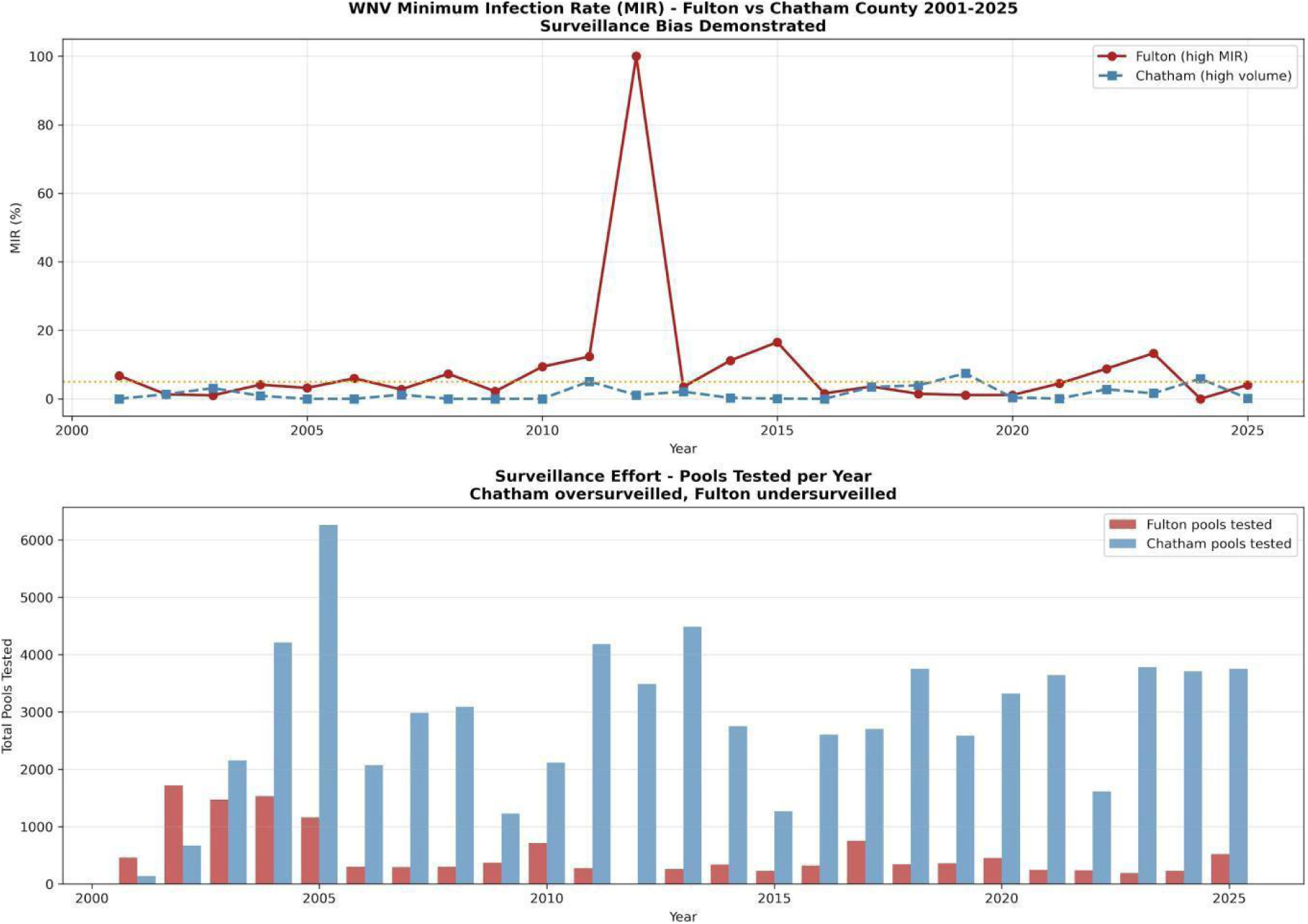
Comparative WNV transmission dynamics between Fulton County and Chatham County (2001–2025). Upper panel: annual MIR comparison. Lower panel: surveillance effort (total pools tested) by year.

**Figure 6:**
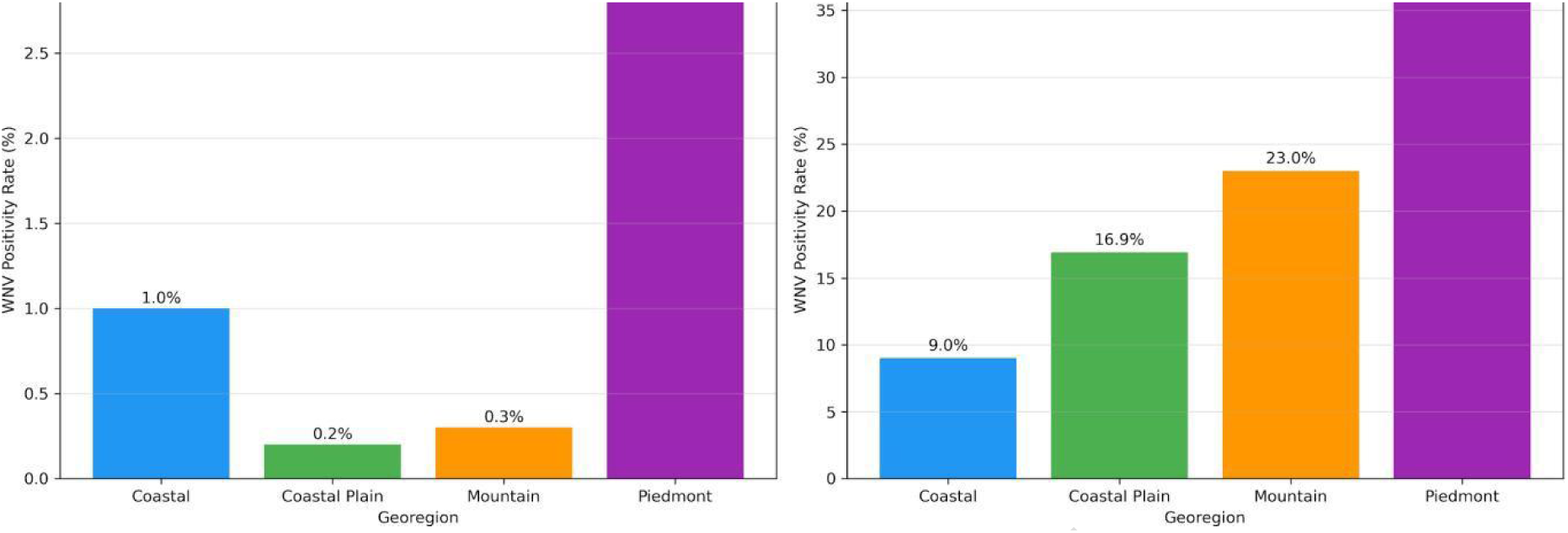
WNV positivity rates by ecological region in Georgia, comparing mosquito pool infection rates (left panel) and dead bird positivity (right panel) across Coastal Plains, Piedmont, Mountain, and Coastal regions.

**Figure 7:**
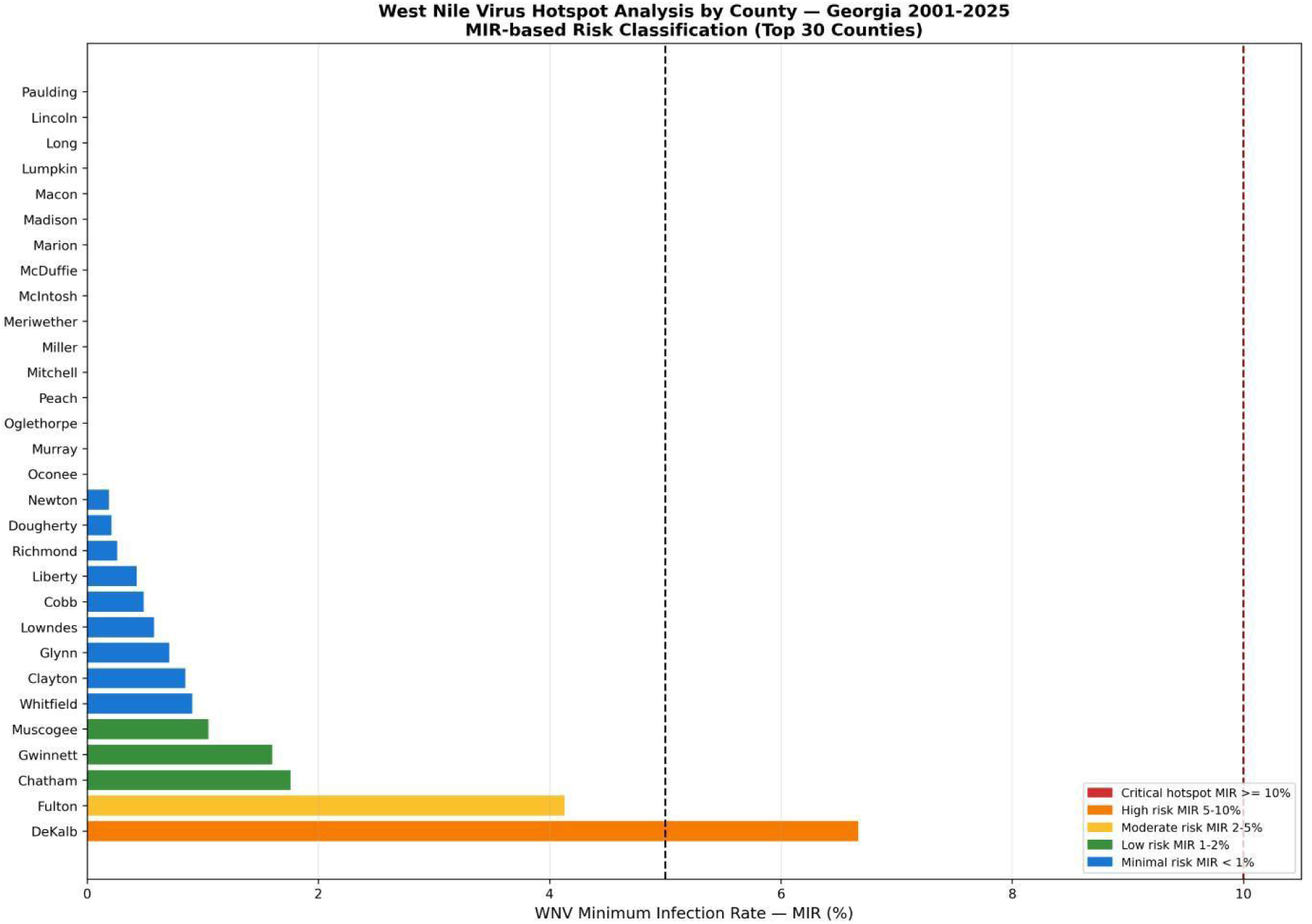
County-level WNV risk classification based on MIR for the top 30 Georgia counties (2001–2025). Color coding: red = critical hotspot (MIR ≥10%), orange = high risk (5–10%), yellow = moderate (2–5%), green = low (<2%).

Highly localized predictive indices successfully delineated this divergence, as WNV risk probabilities approached 0.95–1.00 in the highly urbanized Atlanta metropolitan corridor (DeKalb, Fulton, Cobb, and Gwinnett counties) and Savannah (Chatham County), whereas EEEV risk probabilities peaked at 0.85–0.98 in the rural, swamp-dense coastal plain counties of South Georgia (e.g., Clinch, Ware, and Charlton counties), dropping to near-zero (<0.05) in the northern Piedmont region. In contrast, Chatham County, the most intensively monitored county in the datasets, comprising 72,582 tested pools, recorded a MIR of only

### Equine Arboviral Cases and Clinical Outcomes

A total of 651 equine neurological cases were documented, comprising 356 of confirmed WNV cases (54.7%) and 286 EEEV-confirmed cases (43.9%). Notwithstanding these higher total case counts, WNV exhibited epidemic dominance in only 8 of 25 study years, characterized by four explosive transmission peaks. EEEV demonstrated subtle presence in 17 of 25 years indicating persistent endemic circulation rather than epidemic amplification, a fundamentally distinct transmission dynamics with significant implications for equine vaccination policy. This underscores the critical importance of systematic vaccination as a One Health intervention linking equine and human arboviral disease prevention.

### Climate-Transmission Correlation

A positive correlation was observed between annual WNV MIR and temperature anomalies over the 25-year study period (**Figure 8**). Although moderate, this correlation is consistent with established associations between elevated ambient temperatures and accelerated *Culex* mosquito development rates, increased blood-feeding frequency, and a reduced extrinsic incubation period for WNV (LaDeau et al 2007). The four major transmission peaks in 2001, 2011, 2017–2019, and 2024 coincided with years of above-average temperature anomalies in Georgia, supporting the hypothesis that climate variability is a significant driver of WNV transmission dynamics. Temperature anomalies in Georgia especially in the Piedmont region have an out-sized influence on West Nile virus (WNV) transmission because they amplify every ecological component of the mosquito–bird–virus system. Georgia’s climate already trends warm and humid, but when seasonal temperatures rise above long-term averages, even by 1–3°C, the shift rapidly accelerates *Culex quinquefasciatus* development in urbanized areas like Atlanta where storm-drain networks and dense neighborhoods create ideal larval habitats. This warmer than normal range triggers earlier mosquito emergence, while anomalously hot summers will shorten the extrinsic incubation period of WNV inside the mosquito, allowing vectors to become infectious sooner. These anomalies also intensify urban heat island effects, particularly in metro Atlanta, where night time temperatures remain elevated and extend the seasonal window for viral amplification in birds and subsequent spillover to humans (Scott and Weaver 1989, Turell et al 2005,Stone et al 2010) .

**Figure 8:**
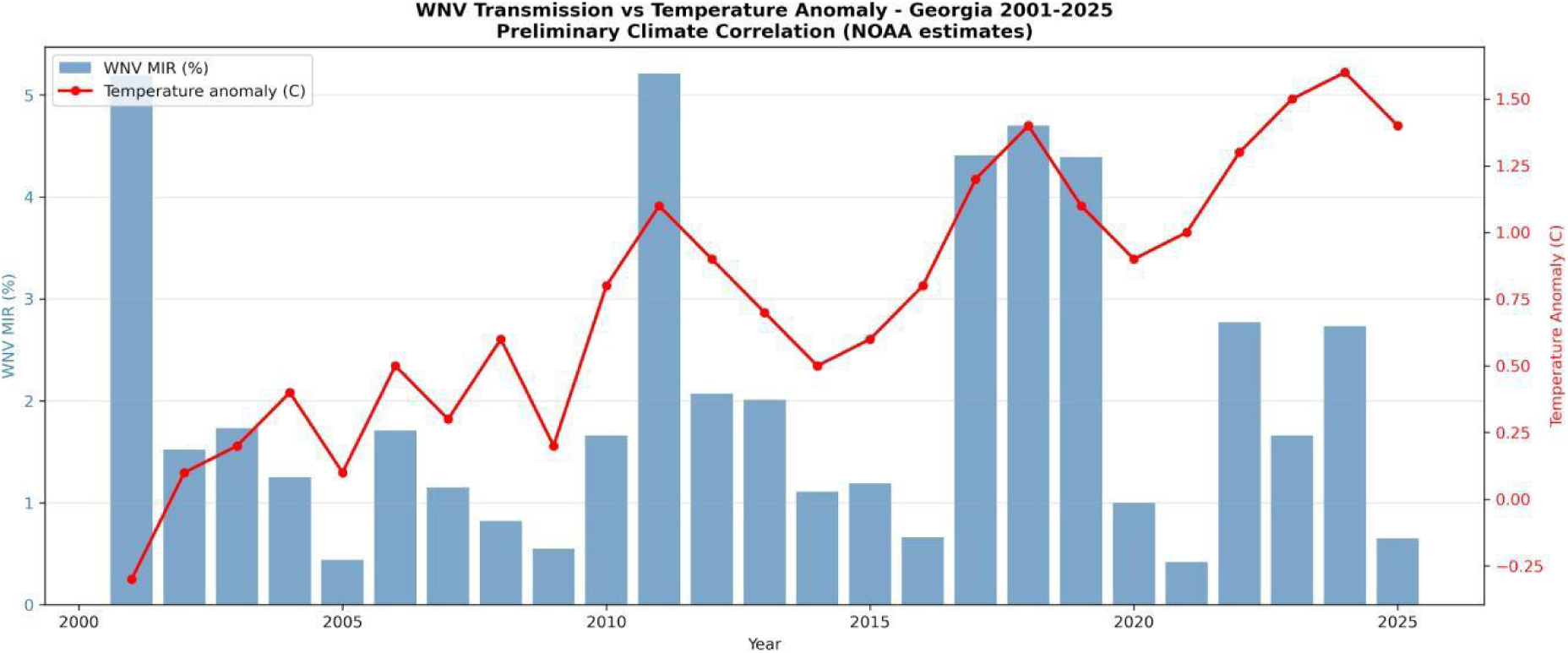
Correlation between annual WNV Minimum Infection Rate and temperature anomalies in Georgia (2001–2025). Bars represent MIR; line represents temperature surges.

## Discussion

This study presents a comprehensive multi-host spatio-temporal assessment of mosquito and arboviral surveillance data conducted in the state of Georgia. The data represent 160,294 mosquito pool records, 7,498 avian mortality reports, and 651 equine neuroinvasive cases documented across a 24-year time-frame. Our findings contribute to advancing understanding of WNV and EEEV transmission dynamics across three critical dimensions: temporal cyclicity, vector competence, and climate-driven transmission dynamics.

The identification of an approximately seven-year periodicity in WNV outbreaks, with major peaks in 2011, 2017–2019, and 2024 has significant implications for strategic arboviral outbreak preparedness. If this cyclical pattern holds, the next major amplification of transmission may be anticipated around 2031. This may be due to the fact that following the 2011 peak, WNV cases crashed but exhibited a highly cyclic pattern, with major resurgences in 2017-2019 (∼60 cases) and 2024 (∼70 cases). This pattern reflects a deeper eco-epidemiological rhythm driven by interactions among avian reservoir immunity, *Culex* population dynamics, and climate variability. This mirrors a classic example of avian herd immunity which rises following large outbreaks, suppressing transmission for several years until population turnover, waning immunity, and recruitment of naive juvenile birds restore conditions favorable for viral amplification. This periodicity is reinforced by temperature anomalies, drought cycles, and urban heat island effects, which periodically accelerate mosquito development and shorten the viral extrinsic incubation period (Dohm et al 2002, McMillan et al 2019).

The central finding of this study is the systematic inverse association between mosquito surveillance intensity and true transmission risks in counties misclassified as arboviral cold spots. Chatham County’s intensive monitoring (72,582 pools, MIR=1.76%) has historically dominated the state’s surveillance profile, while DeKalb (MIR=6.67%) and Fulton (MIR=4.13%) counties in the Atlanta Piedmont corridor have received comparatively lower surveillance efforts despite exhibiting markedly higher transmission intensity. Though Georgia is a home rule state, this trend aligns with mosquito surveillance bias and gaps often encountered in other vector control programs, where monitoring infrastructure creates path-dependent resource allocation that may not reflect contemporary ecological risk distributions.

The dominance of the Piedmont ecological region as the principal hot-spot of WNV transmission risk and predictable outbreaks, with MIR values peaking at 4.39% in 2019, is attributable to multiple interacting urban ecological factors and a lack of comprehensive mosquito control efforts. The Atlanta metropolitan area’s urban heat island effect elevates ambient temperatures by 1–3°C compared to adjacent rural regions, accelerating *Culex quinquefasciatus* development and prolonging transmission seasons (Curren et al 2017). Extensive impervious surface coverage may help to redirect storm water into catch basins and drainage infrastructure which seamlessly provide optimal larval habitat for *Cx. quinquefasciatus,* which can further amplify enzootic transmission cycles (Rueda et al 1990; Rosenberg et al 2018,).

The contrasting transmission dynamics of WNV and EEEV identified through equine case analysis merit particular attention. EEEV predominated in 17 out of 25 study years, despite lower total case counts recorded, indicating persistent low-level enzootic circulation maintained by *Culiseta melanura* within avian reservoirs in Georgia’s wetland habitats. This endemic pattern differs fundamentally from WNV’s epidemic amplification dynamics and suggests that EEEV risk management requires a tailored mosquito surveillance and intervention approach for *Culiseta melanura* and other bridge vectors such as *Coquillettidia perturbans, Culex nigripalpus and Culex quinquefasciatus*.

The positive climate-transmission correlation observed in this analysis, though moderate, is consistent with the growing body of evidence associating climatic anomalies, especially temperature and precipitation, to both WNV and EEEV transmission amplification (Armstrong and Andreadis 2010, Andreadis et al 2008; 2014). The moderate correlation coefficient likely captures the multi-factorial nature of WNV dynamics, wherein temperature interacts with precipitation regimes, land use change, wild bird migration phenology, and variability in mosquito surveillance effort to produce the observed inter-annual MIR fluctuations. Disentangling these interactions through multivariate climate modelling can support future mosquito surveillance control strategies in Georgia.

## Conclusion

This 25-year multi-host surveillance analysis identifies Dekalb and Fulton counties as emerging persistent WNV transmission hot-spots in Georgia. Unfortunately, these counties are systematically underrepresented within local mosquito surveillance infrastructure due to limited funding. The seven-year epidemic cyclic patterns observed in the data set, in conjunction with the positive climate-transmission correlation, establishes a predictive basis for anticipating the next major WNV outbreak around 2031, prompting geographically targeted public health interventions. Prioritizing reallocation of surveillance and mosquito control resources toward DeKalb and Fulton counties, as well as incorporating climate variables into early warning systems, and adoption of a comprehensive One Health surveillance approach are recommended priorities for Georgia’s public health infrastructure. Our findings demonstrate a clear ecological divergence, depicting WNV as an urban, drought-amplified arboviral disease, centered around metropolitan hubs, while EEEV is a rural, wetland-dependent arboviral disease restricted to the acidic swampy habitats of Georgia. These results aim to support public health agencies with localized, predictive patterns to guide re-direction of proactive surveillance of mosquito vectors and their associated arboviruses in emerging WNV and EEEV hot spots in Georgia.

## Acknowledgements

The authors would like to thank the Georgia Department of Public Health (GDPH), Chatham County Mosquito Control Program, Fulton Health Department, DeKalb Health Departments, the Black Fly Research and Resource Center, and the Department of Entomology, University of Georgia.

## Ethics Statement

The study does not require any ethical approval.

## Conflicts of Interest

The authors declare no conflict of interest

## Data Availability

All data produced in the present work are available upon request from the Georgia Department of Public Health (GDPH).

## Notes

### Competing Interest Statement

The authors have declared no competing interest.

### Summary of Updates

This version has been revised to update the authorship arrangement only. (THUY VI THI NGUYEN 1, BRIDGET O. AITO-BOBADOYE 2, ADEM FILALI 3 , ELMER W. GRAY 2, SKYLER M. KERR 2 , ROSMARIE KELLY 1.)

